# Local Anesthetic Systemic Toxicity in Pregnancy: A Retrospective Cohort Analysis

**DOI:** 10.1101/2024.02.25.24303333

**Authors:** Micah K. de Valle, Michael Adkison, Cooper Stevenson, Ruhi Maredia, Shobana Murugan

**Author notes:** **Address correspondence to:** Miss Micah K. de Valle, BS, RN, John Sealy School of Medicine, University of Texas Medical Branch, 301 University Blvd, Galveston, TX. **Funding:** The authors declare no sources of funding for this manuscript.

## Abstract

**Introduction:** Local Anesthetic Systemic Toxicity (LAST) is a rare complication of regional anesthesia. Pregnancy is a risk factor due to gestational physiologic changes. Labor and disorders of pregnancy can mask or delay symptoms of LAST, slowing appropriate intervention. This study examines LAST within a larger cohort and identifies features that help distinguish LAST in pregnancy from nonpregnant patients.

**Methods:** The TriNetX database was used to compare pregnant and nonpregnant patients with LAST from 2013 to 2023. Cohorts were matched on age, race, obesity status, diabetes, metabolic disorders, local anesthetic type, and cardiovascular, liver, kidney, and respiratory disease. Outcomes included prodromal symptoms of LAST and symptoms of cardiac and central nervous system excitation and depression.

**Results:** Matching occurred for 276 pregnant and 276 nonpregnant patients. Pregnant cohorts had a significantly higher risk of cardiac depression (RR, 1.96 [95% CI 1.44 - 2.66], p<0.01) and significantly lower risk of cardiac excitation (RR, 0.38 [95% CI 0.22-0.63], p<0.01), prodromal symptoms (RR, 0.17 [95% CI 0.09 - 0.33], p<0.01), central nervous system excitation (RR, 0.44 [95% CI 0.21-0.90], p=0.02), and central nervous system depression (RR, 0.24 [95% CI 0.13-0.48], p<0.01) than nonpregnant cohorts.

**Conclusion:** Pregnant patients with LAST were more likely to exhibit cardiac depression and less likely to manifest prodromal symptoms, cardiac excitation, and central nervous system excitation and depression than nonpregnant patients. Physiological changes during pregnancy and prompt detection and treatment may explain these differences. These findings highlight the variable nature of LAST and how pregnancy may influence its clinical presentation.

**Key Messages:** Local anesthetic systemic toxicity, a rare complication of regional anesthesia, has variable manifestations within the cardiovascular and central nervous systems. This study identifies local anesthetic systemic toxicity in a larger cohort relative to previous literature, revealing a distinct presentation in pregnant patients compared to their nonpregnant counterparts. These findings emphasize the diverse nature of local anesthetic systemic toxicity and indicate severe complications in pregnancy. Recognition of subtle clinical manifestations in pregnant patients receiving local anesthesia aids accurate diagnosis and timely intervention in the event of toxicity. The present study provides additional insight into local anesthetic systemic toxicity and sets the stage for further investigations in pregnant patients.

## INTRODUCTION

### Background

Local Anesthetic Systemic Toxicity (LAST) is a rare but potentially life-threatening complication of local anesthesia. LAST occurs when increased plasma levels of local anesthetic alter cardiovascular (CV) and central nervous system (CNS) voltage-gated ion channels.^1^ While LAST classically manifests with CNS and CV excitation followed by depression, presentation is highly variable.^1^ This variability, with its rarity, contributes to under-recognition, misdiagnosis, and challenging management.^1,2^ Estimated incident rates hover around 1-2 per 1000 nerve blocks; however, the true incidence of LAST remains uncertain due to limited research and diagnostic ambiguity.^3,4^

Factors influencing local anesthetic absorption include dosage, injection site perfusion, and drug clearance.^1,5,6^ Thus, exceeding recommended dosages or accidental intravascular injection increases the risk of LAST. Patient characteristics like extremes of age or weight and comorbidities affecting the heart, liver, and kidneys may increase LAST susceptibility by impacting absorption and clearance.^1,2^

### LAST in Pregnancy

Pregnancy is another risk factor that may precipitate or exacerbate LAST.^1,7^ Labor and diseases of pregnancy can mimic, mask, or slow symptom onset, potentially complicating management by preventing diagnosis and delaying treatment.^1^ Physiological changes during gestation impact local anesthetic dynamics, including absorption, peak free plasma concentration, clearance, and intravascular injection likelihood.^1,3,5^ Increased cardiac output and blood volume can enhance injection site perfusion, accelerating anesthetic absorption.^1,3,5^ These changes, coupled with limited space from the fetus, facilitate venous engorgement in the epidural space, resulting in increased absorption and catheter migration risk in neuraxial procedures.^1,3,5^ Furthermore, pregnancy-related reduction in alpha-1-glycoproteins, which bind free local anesthetic, raises plasma levels of unbound anesthetic, allowing toxic accumulation within tissues.^1,5^

The risk of LAST in pregnancy necessitates careful consideration by clinicians, as local anesthesia is frequently used in the inpatient peripartum setting.^1^ Common applications include neuraxial blocks for labor analgesia or cesarean deliveries.^1^ Additional indications include pudendal, paracervical, or transabdominal plane blocks, perineal laceration repairs, and outpatient procedures.^1^ Bupivacaine and lidocaine are the most commonly used local anesthetics in this population.^1^ Despite widespread use in labor epidural analgesia, bupivacaine can increase LAST risk. Its vasodilatory and lipophilic properties expedite local anesthetic absorption and promote cardiac toxicity, respectively.^1,5^ Cardiomyocyte irritability is further potentiated by estrogen and progesterone fluctuations during pregnancy, increasing susceptibility to tachyarrhythmias related to LAST.^1^ Finally, risk factors within pregnancy, such as advanced maternal age and preeclampsia, are associated with impaired local anesthetic clearance by altering renal and liver function.^1,5^

The risks described introduce uncertainty regarding safe local anesthetic dosing in pregnant patients.^1,6^ The fetus contributes further complexity, as one report demonstrated newborn toxicity from maternal lidocaine intoxication.^1,8^ Despite these considerations, existing studies on LAST in pregnancy are primarily limited to review articles and case reports due to ethical concerns of randomized controlled trials.^1^

### Objective

This study seeks to identify LAST within a larger population cohort, distinguish unique features of LAST in pregnant patients versus their nonpregnant counterparts, and contextualize findings relative to existing literature.

This article’s content was previously presented as a poster at the 64^th^ Annual National Student Research Forum at The University of Texas Medical Branch on April 1, 2023.

## METHODS

### Data Source

We conducted a retrospective propensity score-matched cohort study utilizing the TriNetX US Collaborative Network. This network contains over 90.9 million patients derived from electronic medical records (EMR) in 56 different healthcare organizations (HCOs) across the United States. TriNetX provides aggregated health counts and statistical summaries of deidentified patient data but does not disclose Protected Health Information, personal data, or the ability to connect patient data to specific HCOs within the network. Given these safeguards, this study is exempt from approval by the Institutional Review Board.

### Study Population

We identified two distinct populations of interest as pregnant (P+/LAST+) and nonpregnant (P-/LAST+) female patients aged 18-45 who experienced an incident of LAST after administration of lidocaine, bupivacaine, or ropivacaine within the timeframe of January 2013 to January 2023. All male patients were excluded from the study before cohort identification. Pregnancy was defined as having one or more ICD-10-CM diagnostic codes (Z33, Z34, Z3A, O00-O9A) or ICD-10-PCS procedure code (10). LAST was defined using three ICD-10 codes (T41.3X4, T41.3X5, T41.3X1) described in previous literature.^9^ The timing of LAST events was specified to coincide with the pregnant or nonpregnant status of the respective cohorts.

### Covariates

To ensure comparability between pregnant and nonpregnant cohorts, we employed propensity score matching utilizing established risk factors for LAST. Cohort matching included patient demographics, comorbidities affecting the likelihood of pregnancy, and potential confounders influencing our outcomes, ensuring observed differences were attributed to pregnancy. TriNetX could not specify the point at which the two groups diverged (i.e. when one cohort became pregnant). To address this, we implemented matching from 10 years up to nine months before the LAST event, ensuring uniformity in nonpregnancy status and other matched characteristics until the nine-month time point, with half experiencing pregnancy thereafter.

Cohorts were matched on age at the LAST event, race, and previous encounters involving the administration of lidocaine, bupivacaine, or ropivacaine. Matching also extended to the following diagnoses: overweight and obesity, diabetes mellitus, ischemic heart disease, other heart diseases, hypertensive disease, tobacco and nicotine use, liver disease, chronic kidney disease, chronic lower respiratory disease, and disorders of lipoprotein metabolism. Anesthetic allergies, mitochondrial metabolic disorders, and carnitine deficiencies were initially considered but excluded due to the absence of documented cases in either cohort.

TriNetX extrapolates laboratory data from any point within our established 10-year matching period. To mitigate confounding biases, unless explicitly stated, all laboratory values represented the most recent values within the EMR during the study time frame. Age was stratified into 18-31 and 32-45-year-olds.

In total, 20 different characteristics were matched in this study.

### Outcomes

The outcomes were tabulated within a time frame ranging from the day of the LAST event to one day afterward. This assessment aimed to account for the acute nature of the event, capture the disease’s progression during this period, and minimize outcome variations attributable to other causes.

Evaluated outcomes included prodromal symptoms and symptoms of CV excitation, CNS excitation, CV depression, and CNS depression. Specific outcomes within each of the five categories are as follows: prodromal symptoms (tinnitus, disturbances of skin sensation, disorientation unspecified, altered mental status unspecified, disturbances of smell and taste, auditory hallucinations, other abnormal auditory perceptions, visual disturbances, visual hallucinations, hallucinations unspecified, slurred speech, dizziness and giddiness), cardiac excitation (tachycardia, cardiac arrhythmias, paroxysmal tachycardia, atrial fibrillation and flutter, hypertensive crisis, elevated blood-pressure reading without a diagnosis of hypertension, palpitations, secondary hypertension unspecified, other secondary hypertension, dyspnea, chest pain), cardiac depression (atrioventricular and left bundle-branch block, bradycardia, cardiogenic shock, cardiac arrest, hypotension due to drugs, hypotension unspecified, other hypotension, syncope and collapse, conduction disorders), CNS excitation (muscle spasm, epileptic seizures related to external causes, tremor unspecified, abnormal involuntary movements, abnormal head movements, abnormal reflex, convulsions, myoclonus, fasciculation, dysarthria and anarthria, restlessness and agitation), and CNS depression (somnolence, stupor and coma, respiratory arrest, pulmonary collapse, hypoxemia, apnea, acute respiratory failure). Vital sign results identifying respiratory rates ≤ 11 breaths per minute, indicative of bradypnea, were also included under the CNS depression category. Corresponding ICD-10-CM diagnosis codes for all outcomes within the five categories are displayed in Appendix A.

### Statistical Analysis

We employed a 1:1 propensity score-matching approach using greedy nearest neighbor matching to establish balanced cohorts with matched baseline characteristics from up to 10 years to nine months preceding the LAST event. This ensures that each patient in one group is precisely matched with one patient in the other by pairing them with their nearest neighbor based on propensity scores. A caliper distance of 0.1 pooled standard deviations of the logit of the propensity score was applied to ensure equilibrium across cohorts. As such, matches were only considered valid if the absolute difference in propensity scores between matched individuals was within this threshold. Characteristics falling outside of the specified time range were excluded from the study. Well-matched characteristics were defined as those with a p-value value ≥ 0.05. Assessment of cohort outcome associations utilized risk ratio analysis with 95% confidence intervals. Statistical significance was determined by a *p*-value ≤0.05.

## RESULTS

### Study Population

The database identified 839 pregnant patients with an incidence of LAST (P+/LAST+) and 305 nonpregnant patients with an incidence of LAST (P-/LAST+) for pre-propensity score matching analysis. Both cohorts were exclusively female. Pregnant cohorts were selected from 35 different HCOs, with the following geographic distribution across the United States: northeast (16%), mid-west (41%), south (34%), west (7%), and unknown (1%). Nonpregnant cohorts were selected from 40 different HCOs, with the following geographic distribution across the United States: northeast (29%), mid-west (13%), south (35%), west (20%), and unknown (3%). Before matching, the mean age at the time of the LAST incident was 30.1 ± 5.6 years for P+/LAST+ and 34.9 ± 7.4 for P-/LAST+ cohorts (p<0.01). Before matching, the P+/LAST+ cohort contained predominantly White (60.7%) and Black/African American (20.4%) patients. The P-/LAST+ cohort was also predominantly White (68.9%) and Black/African American (14.8%) before matching.

### Matching Analysis

Post-matching, 276 patients remained in each cohort. Since mean age during the LAST diagnosis significantly differed after matching (p<0.01), age was stratified into two ranges for matching to minimize confounders due to age differences, as described above. Following this modification, the post-match analysis revealed no significant differences in the distribution of patients across each age or race category.

Before matching, the P-/LAST+ cohort had higher percentages of patients with chronic kidney disease, liver disease, heart disease, diabetes mellitus, and hypertensive disease than the P+/LAST+ cohort (all p<0.01). No significant differences between cohorts were observed post-matching. Additionally, there were no significant differences in local anesthetic type before or after matching. Complete demographics before and after matching are summarized in Table 1.

**Table 1:** Demographics, Diagnoses, and Local Anesthetics Before and After Propensity Score Matching.

|  | Before Propensity Score Matching |  |  | After Propensity Score Matching |  |  |
| --- | --- | --- | --- | --- | --- | --- |
|  | *Pregnant*<br>(n=839) | *Nonpregnant*<br>(n=305) | *p-value*<sup>a</sup> | *Pregnant*<br>(n=276) | *Nonpregnant*<br>(n=276) | *p-value*<sup>b</sup> |
| Age at Index, mean ± sd | 30.1 ± 5.6 | 34.9 ± 7.4 | <0.01 | 32.8 ± 5.0 | 34.5 ± 7.3 | <0.01 |
| 18-31, (%) | 493 (58.8) | 86 (28.2) | <0.01 | 82 (29.7) | 82 (29.7) | 1 |
| 32-45, (%) | 346 (41.2) | 219 (71.8) | <0.01 | 194 (70.3) | 194 (70.3) | 1 |
| Race, (%) |  |  |  |  |  |  |
| White | 509 (60.7) | 210 (68.9) | 0.01 | 203 (73.6) | 198 (71.7) | 0.63 |
| Black or African American | 171 (20.4) | 45 (14.8) | 0.03 | 32 (11.6) | 34 (12.3) | 0.79 |
| Asian | 47 (5.6) | 10 (3.3) | 0.11 | 10 (3.6) | 10 (3.6) | 1 |
| American Indian or Alaska Native | 10 (1.2) | 10 (3.3) | 0.02 | 10 (3.6) | 10 (3.6) | 1 |
| Native Hawaiian or Other Pacific Islander | 10 (1.2%) | 10 (3.3) | 0.02 | 10 (3.6) | 10 (3.6) | 1 |
| Diagnoses, (%) |  |  |  |  |  |  |
| Overweight and Obesity | 109 (13) | 42 (13.8) | 0.73 | 29 (10.5) | 30 (10.9) | 0.89 |
| Diabetes Mellitus | 21 (2.5) | 19 (6.2) | <0.01 | 10 (3.6) | 10 (3.6) | 1 |
| Ischemic Heart Disease | 10 (1.2) | 13 (4.3) | <0.01 | 10 (3.6) | 10 (3.6) | 1 |
| Heart Disease, Other | 34 (4.1) | 38 (12.5) | <0.01 | 12 (4.3) | 19 (6.9) | 0.20 |
| *Hypertensive Disease* | 44 (5.2) | 46 (15.1) | <0.01 | 23 (8.3) | 23 (8.3) | 1 |
| *Nicotine Dependence* | 60 (7.2) | 18 (5.9) | 0.46 | 14 (5.1) | 15 (5.4) | 0.85 |
| *History of Nicotine Dependence* | 39 (4.6) | 19 (6.2) | 0.28 | 14 (5.1) | 15 (5.4) | 0.85 |
| *Liver Disease* | 15 (1.8) | 18 (5.9) | <0.01 | 10 (3.6) | 12 (4.3) | 0.60 |
| *Chronic Kidney Disease* | 10 (1.2) | 12 (3.9) | <0.01 | 10 (3.6) | 10 (3.6) | 1 |
| *Chronic Respiratory Disease* | 102 (12.2) | 47 (15.4) | 0.15 | 36 (13.0) | 38 (13.8) | 0.80 |
| *Disorders of lipoprotein metabolism* | 28 (3.3) | 15 (4.9) | 0.21 | 10 (3.6) | 10 (3.6) | 1 |
| Local Anesthetics, (%) |  |  |  |  |  |  |
| *Lidocaine* | 237 (28.2) | 102 (33.4) | 0.09 | 63 (22.8) | 78 (28.3) | 0.14 |
| *Bupivacaine* | 153 (18.2) | 69 (22.6) | 0.10 | 48 (17.4) | 52 (18.8) | 0.66 |
| *Ropivacaine* | 39 (4.6) | 11 (3.6) | 0.45 | 10 (3.6) | 10 (3.6) | 1 |

Abbreviations: sd, standard deviation
<sup>a,b</sup>: p-value compares characteristics between pregnant and nonpregnant cohorts before propensity score matching<sup>a</sup> and after propensity score matching<sup>b</sup>.

### Clinical Outcomes Analysis

Post-matching relative risk analysis of clinical presentation outcomes revealed the P+/LAST+ cohort had a significantly higher incidence of CV depression (RR, 1.96 [95% CI, 1.44-2.66], p<0.01) than the P-/LAST+ cohort. Additionally, P+/LAST+ cohorts had significantly lower incidences of prodromal symptoms (RR, 0.17 [95% CI 0.09-0.33], p<0.01), CNS excitation (RR, 0.44 [95% CI 0.21-0.90], p=0.02), CNS depression (RR, 0.24 [95% CI 0.13-0.48], p<0.01), and CV excitation (RR, 0.38 [95% CI 0.22-0.63], P<0.01) compared to the P-/LAST+ cohort. Findings are summarized in Table 2. A percentage of each cohort did not return any of the LAST symptoms tested in our analysis, which are listed in Appendix A. Attempts to mitigate the percentage of missing data included a thorough review of LAST manifestations from reliable literature sources to capture all commonly documented symptoms.^2,3,5–7,10–12^ Despite these attempts, patients with unidentified or undocumented symptoms represented ≤48.6% of the P+/LAST+ cohort and ≤21% of the P-/LAST+ cohort.

**Table 2:** Post-Match Relative Risk Analysis.

| Clinical Presentation | *Pregnant*<br>(*n*=276) | *Nonpregnant*<br>(*n*=276) | *RR (95% CI)* | *p-value*<sup>a</sup> |
| --- | --- | --- | --- | --- |
| Prodromal Symptoms | $\leq 10$ | 58 | 0.17 (0.09-0.33) | <0.01 |
| Cardiovascular Excitation | 18 | 48 | 0.38 (0.22-0.63) | <0.01 |
| Cardiovascular Depression | 94 | 48 | 1.96 (1.44-2.66) | <0.01 |
| Central Nervous System<br>Excitation | $\leq 10$ | 23 | 0.44 (0.21-0.90) | 0.02 |
| Central Nervous System<br>Depression | $\leq 10$ | 41 | 0.24 (0.13-0.48) | <0.01 |

Abbreviations: RR, relative risk
<sup>a</sup>: p-value compares clinical presentation outcomes between pregnant and nonpregnant cohorts after propensity score matching<sup>a</sup>.

## DISCUSSION

### Primary Findings

The present study found that both pregnant and nonpregnant cohorts exhibited conventional clinical manifestations of LAST, with varying severity and presentation distribution. Our P+/LAST+ cohort demonstrated a significantly higher incidence of cardiovascular depression, affecting 34.1%, and a lower incidence of cardiovascular excitation (6.52%), prodromal symptoms (≤3.62%), excitatory (≤3.62%), and depressive (≤3.62%) CNS manifestations, than our P-/LAST+ cohort.

Physiologic changes in pregnancy and anesthetic side effects may partly explain observed risk differences in the development of hypotension between our cohorts. During late gestation, a decrease in systemic vascular resistance due to aortocaval compression can cause a decrease in blood pressure.^10–12^ Furthermore, neuraxial blocks can induce hypotension by suppressing the sympathetic nervous system.^13^ Additionally, vigilant cardiovascular monitoring in pregnant patients may lead to earlier detection and intervention. Consequently, this can alter the course of clinical sequelae observed, yielding higher numbers of CV depressive symptoms reported in our pregnant cohort versus their nonpregnant counterparts. Moreover, studies suggest that increased progesterone levels in pregnancy can provide a protective effect to the mother and fetus via nervous system inhibition.^14–16^ This effect may have prevented the manifestation of detectable CNS symptoms in our pregnant cohort. Furthermore, limited prodromal symptoms in our pregnant cohort may be attributed to the ability of labor and pregnancy-related conditions to mask, mimic, or slow the onset of LAST symptoms.^1,2,5^ It may also be attributed to the frequent misdiagnosis and underreporting of LAST, especially in pregnant patients.^1,2,5^ This narrowed symptom distribution in our pregnant cohort suggests that LAST may present unconventionally in pregnancy, likely impeding diagnosis.

### Classic Presentation Versus Reality

Comparing the typical clinical sequence of LAST within the general population to cases of pregnant patients in existing literature reveals notable deviations. Classically, LAST initially manifests with mild prodromal symptoms such as dizziness, disorientation, dysgeusia, tinnitus, and circumoral paresthesia, followed by excitatory CNS and CV symptoms like seizures and tachyarrhythmias, respectively.^5,7,10–12^ Subsequent progression to depressive symptoms ensues with rapid CV and CNS deterioration.^7^ The characteristic clinical sequelae of LAST are summarized in Figure 1. ^5,7,10–12^

**Figure 1:**
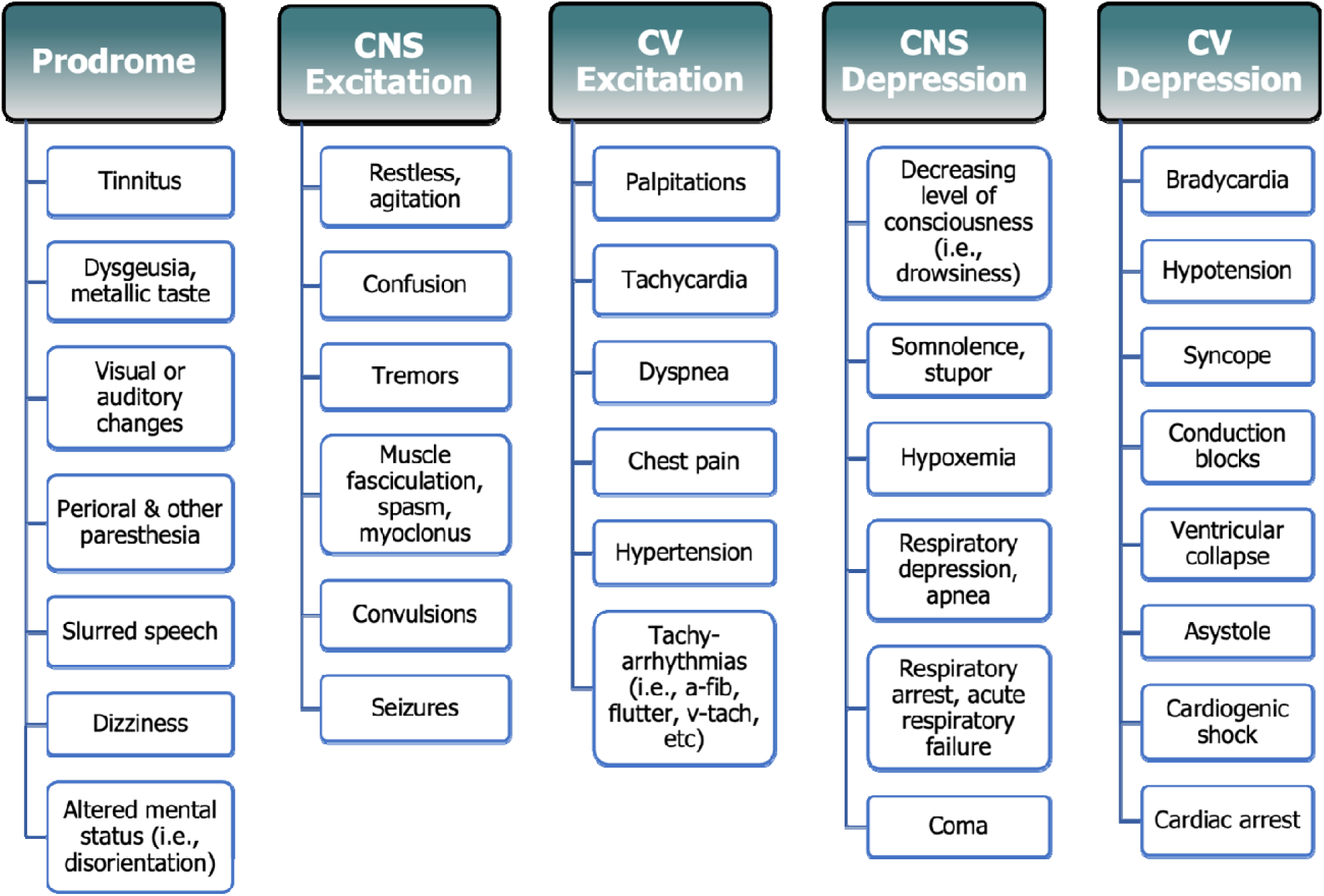
Clinical Presentation of Local Anesthetic Systemic Toxicity. LAST signs and symptoms in classic order of presentation, including prodromal, central nervous system (CNS) excitation, cardiovascular (CV) excitation, CNS depression, and CV depression.

However, many pregnancy reports do not reflect this anticipated clinical sequence, posing diagnostic challenges for clinicians to accurately recognize LAST.^12,17^ For instance, a 29-year-old primigravida at 39 weeks gestation exhibited prodromal symptoms (tinnitus and metallic taste) followed by palpitations, tachycardia, and feelings of dissociation within 50 minutes of receiving combined spinal-epidural analgesia with bupivacaine.^18^ Another case details a term parturient who developed progressive hypotension, nausea, drowsiness, tremors, nystagmus, and loss of consciousness five minutes after receiving an epidural analgesia mixture of levobupivacaine, sufentanil, and clonidine.^19^ Lastly, a severe case recounts a 39-year-old parturient who received ropivacaine for epidural anesthesia and subsequently suffered a seizure, chest pain, pulseless ventricular tachycardia, and cardiac arrest within 22 minutes.^14^

### Relevance and Implications

Amidst this diverse array of clinical sequelae, the present study revealed a lower incidence of prodromal symptoms in pregnancy, affecting ≤6.52% of our pregnant cohort, with none experiencing tinnitus, taste disturbance, disorientation, restlessness or agitation, or tremors. To our knowledge, this finding is consistent with previous literature, as only two reports detail a clear manifestation of prodromal symptoms in pregnant patients.^18,20^ In contrast, 21% of our nonpregnant cohort were found to have prodromal symptoms, which is comparable to other analyses that estimated prodromal symptoms in the general population.^3,11^ These findings suggest that detecting LAST in pregnant patients may be less distinguishable, partly due to absent or mildly presenting prodromes. Moreover, roughly 48.6% or less of our pregnant cohort had no identified symptoms compared to only 21% or less of our nonpregnant cohort. This may be due to the underreporting of symptoms or the inability to distinguish LAST presentations from physiologic and pathologic processes in pregnancy.^1^

Importantly, 34.1% of our pregnant cohort experienced CV depression, primarily attributed to medication-induced hypotension (30% cases), while ≤3.6% exhibited excitatory manifestations like hypertension, palpitations, tachycardia, arrhythmias, seizures, or convulsions. The emergence of cardiovascular depression following the administration of local anesthetics may serve as a sensitive marker of systemic toxicity in pregnant patients, aiding clinicians in differentiating anesthetic toxicity from other obstetric conditions.

A marginal percentage of pregnant cohorts exhibited evidence of severe LAST symptoms and clinical deterioration, including syncope and collapse (≤3.6% cases), cardiac arrest (≤3.6%), respiratory arrest (≤3.6%), acute respiratory failure (≤3.6%), and somnolence, stupor, or coma (≤3.6%). Existing data reports indicate that when severe manifestations of LAST occur in the general population, many patients initially experience CV depressive symptoms, such as bradycardia and hypotension, before progressing to cardiac arrest.^21^ Although only observed in a marginal percentage of patients, awareness of these subtle symptom changes and the projected clinical sequelae may help clinicians identify cases of rapid deterioration and reduce fatal outcomes.

### Limitations

There are limitations to our study to be considered. Firstly, a significant age difference between cohorts post-matching was observed, leading to possible age-related biases. However, cohorts were stratified by age range to mitigate this discrepancy. Additionally, our study’s retrospective design limits our ability to establish causality of reported outcomes. Although we support our findings with existing literature, this underlines the need for further prospective investigations. Further limitation related to our retrospective approach includes a percentage of each cohort (48.6% in P+/LAST+, 21% in P-/LAST+) having a LAST diagnosis without any reported symptoms. This observation is suspected to arise from providers documenting a LAST diagnosis but underreporting actual symptoms within the EMR or because symptoms were not included in our outcomes. Additionally, the reporting practice of TriNetX introduced some ambiguity by presenting outcomes that included any number of patients from one to ten as “10”. We specifically indicated these cases by the patient count, “≤10,” to ensure clarity in our reporting. However, this limits our ability to provide more definite values for those outcomes. Finally, the external validity of the present study is limited by our focus on pregnant and nonpregnant females aged 18-45 in the United States. However, within this demographic, the external validity is supported by the extensive TriNetX database, covering 90.9 million patients from 56 HCOs across various regions in the United States. This integration of real-world clinical data and geographic diversity allows for applicability to multiple healthcare settings and patient populations nationwide.

### Future Directions

Subsequent studies may investigate the specific local anesthetics influencing the observed clinical manifestations, expanding beyond our cumulative focus on lidocaine, bupivacaine, and ropivacaine. Additionally, future investigations should target identifying combined risk factors predisposing pregnant patients to LAST. This exploration can enhance risk stratification and enable the development of proactive management strategies for high-risk patients. Finally, examining the dosage thresholds at which toxicity occurs in pregnant cohorts would be beneficial to aid the establishment of safer dosage guidelines and optimize patient care.

## Conclusion

To our knowledge, this study is the first of its kind to examine LAST in a large cohort of pregnant patients. Our findings indicate that both pregnant and nonpregnant patients who experienced LAST exhibited the typical manifestations of cardiovascular and central nervous system excitation and depression. However, pregnant patients displayed a higher incidence of cardiovascular depression, particularly hypotension. In contrast, nonpregnant patients more often displayed cardiovascular excitation and prodromal, excitatory, and depressive neurologic symptoms.

In conclusion, this study underlines the diverse and rapidly evolving nature of LAST, particularly in pregnant patients. Heightened awareness and vigilance of subtle clinical indications may assist clinicians in promptly and accurately identifing LAST, allowing more timely intervention and minimizing potentially fatal outcomes during local anesthetic administration in pregnant patients.

## Data Availability

All data produced in the present study are available upon reasonable request to the authors.

## ACKNOWLEDGEMENTS

TriNetX

### Appendix A

**Table 8:** Corresponding Codes to Outcomes Tested.

| **Outcomes** | **Code** |
| --- | --- |
| **Prodromal Symptoms** | **ICD-10 Codes** |
| Tinnitus | H93.1 |
| Disturbances of skin sensation | R20 |
| Disorientation | R41.0 |
| Altered mental status | R41.82 |
| Disturbances of smell and taste | R43 |
| Abnormal auditory perceptions | H93.2 |
| Dizziness and giddiness | R42 |
| Visual disturbances | H53 |
| Slurred speech | R47.81 |
| Auditory hallucinations | R44.0 |
| Visual hallucinations | R44.1 |
| Hallucinations, unspecified | R44.3 |
| **Cardiovascular Excitation** | **ICD-10 Codes** |
| Tachycardia | R00.0 |
| Cardiac arrhythmias | I49 |
| Paroxysmal tachycardia | I47 |
| Atrial fibrillation and flutter | I48 |
| Hypertensive crisis | I16 |
| Elevated blood-pressure reading, without diagnosis of hypertension | R03.0 |
| Palpitations | R00.2 |
| Secondary hypertension | I15.8, I15.9 |
| Dyspnea | R06.0 |
| Chest pain | R07.9 |
| **Cardiovascular Depression** | **ICD-10 Codes** |
| Atrioventricular and left bundle-branch block | I44 |
| Bradycardia | R00.1 |
| Cardiogenic shock | R57.0 |
| Cardiac arrest | I46 |
| Hypotension due to drugs | I95.2 |
| Syncope and collapse | R55 |
| Conduction disorders | I45 |
| Other hypotension | I95.8, I95.9 |
| Central Nervous System Excitation | ICD-10 Codes |
| Restless and agitation | R45.1 |
| Muscle spasm | M62.83 |
| Fasciculation | R25.3 |
| Epileptic seizures related to external causes | G40.5 |
| Epilepsy and recurrent seizures | G40 |
| Tremor | R25.1 |
| Abnormal involuntary movements | R25, R25.8 |
| Abnormal head movements | R25.0 |
| Abnormal reflex | R29.2 |
| Convulsions | R56, R56.9 |
| Myoclonus | G25.3 |
| Dysarthria and anarthria | R47.1 |
| Central Nervous System Depression | ICD-10 Codes |
| Somnolence, Stupor, and Coma | R40 |
| Respiratory Arrest | R09.2 |
| Hypoxemia | R09.02 |
| Apnea | R06.81 |
| Acute Respiratory Failure | J96.0 |
| Altered Mental Status, Unspecified | R41.82 |
| Pulmonary collapse | J98.1 |
| Respiratory rate ≤ 12 breaths per minute | TNX:9073 |

